# Mesoscale CISS Imaging for the Detection of Dural Defects in Spinal CSF Leaks

**DOI:** 10.1101/2025.07.14.25331467

**Authors:** Michelle L. Wegscheid, Arindam R. Chatterjee, Cyrus A. Raji, Martin N. Reis, Nicholas P. Fleege, Sherwin N. Azad, John Ogunlade, Ananth K. Vellimana, Manu S. Goyal, Arash Nazeri

**Affiliations:** Mallinckrodt Institute of Radiology, Washington University School of Medicine, St. Louis, MO; Department of Neurosurgery, Washington University School of Medicine, St. Louis, MO; Department of Neurology, Washington University School of Medicine, St. Louis, MO

## Abstract

**Background and Purpose:** Spontaneous intracranial hypotension (SIH) is most often attributable to cerebrospinal fluid (CSF) leaks from the spinal canal, yet precise localization of dural defects remains challenging. While fat-saturated heavily T2-weighted MR myelography (HT2-MRM) is sensitive to spinal longitudinal extradural CSF collections (SLECs), it does not reliably localize the leak site. This study evaluates a two-stage spine MRI protocol incorporating high resolution, mesoscale 3D constructive interference in steady state (meso-CISS) for targeted CSF leak localization.

**Materials and Methods:** Twenty consecutive patients with suspected SIH underwent a standardized total spine MRI protocol, including 3D HT2-MRM for SLEC screening. In cases with positive SLEC or indeterminate findings, meso-CISS was performed for targeted high-resolution leak localization.

**Results:** Eight patients (40%) demonstrated SLECs on HT2-MRM, prompting further evaluation with meso-CISS. Dural defects were successfully identified in five patients (1.5-8 mm in size). In one case, meso-CISS delineated intrathecal protrusion of ligamentum flavum calcification, suggesting an underlying defect. Compared with HT2-MRM, meso-CISS provided superior visualization of dural defects, arachnoid scarring, and neo-membrane formation. Two cases were limited by motion or metal artifacts.

**Conclusion:** A two-stage spine MRI protocol incorporating HT2-MRM and meso-CISS improves the localization of dural defects in SIH. Meso-CISS enhances visualization of small dural tears and associated pathology, facilitating targeted interventions and potentially reducing reliance on invasive myelography. Further studies are warranted to assess its broader clinical impact.

## INTRODUCTION

Spontaneous intracranial hypotension (SIH) is a neurological disorder most often attributable to cerebrospinal fluid (CSF) leakage from the spinal canal. Patients with SIH typically present with orthostatic headaches that worsen upon standing and improve when lying down[1]. Additional symptoms may include nausea, dizziness, tinnitus, and cognitive dysfunction, all of which can significantly impact quality of life[2]. Despite growing awareness, SIH remains underdiagnosed due to its variable clinical presentation and challenges in precisely localizing CSF leaks[3].

Spinal CSF leaks may be described in three major categories, with two associated with spinal longitudinal extradural CSF collections (SLECs): Type 1, caused by dural tears often due to discogenic microspurs or osteophytes, and Type 2, typically representing a lateral dural tear with secondary herniation of arachnoid matter or ruptured meningeal diverticula[4–8]. Precise localization of dural defects is essential for targeted treatment and surgical intervention, particularly when conservative measures fail. Although fat-saturated heavily T2-weighted MR myelography (HT2-MRM) demonstrates high accuracy in detecting SLECs[3, 9], it does not reliably correlate the craniocaudal extent of the SLEC with the exact leak site[10, 11]. Recent studies indicate that indirect findings on spine MRI may aid in localizing CSF leaks[11, 12]; however, precise identification of small dural breaches, as tiny as 1-2 mm, remains a significant challenge and often requires contrast myelography for definitive confirmation[1, 13]. Both dynamic CT myelography (CTM) and digital subtraction myelography (DSM) must capture the full extent of the SLEC to precisely identify the leak site. However, dynamic myelography can be technically challenging and is associated with high radiation doses, especially when repeat examinations are required, which further contributes to patient discomfort. Consequently, there is substantial impetus to develop noninvasive imaging strategies that directly detect dural defects. Such advancements could allow for more targeted myelographic evaluation, reducing radiation exposure and procedure time, or potentially obviating the need for myelography altogether. In addition, CSF leaks may be associated with secondary complications, such as arachnoid scarring and extradural neo-membrane formation[14], that may not be readily identifiable on conventional dynamic myelography[15].

Balanced steady state free-precession (b-SSFP) techniques, such as constructive interference in steady state (CISS), have been used for high resolution spine imaging[16–18], including evaluation of spinal CSF leaks in select contexts such as superficial siderosis and ventral dural defects[19–21]. High resolution CISS imaging allows sharp visualization of fine structures in the CSF due to high signal to noise efficiency, high contrast between CSF and neural tissue and meninges, inherent flow compensation, and steady-state k-space filling[16, 22, 23]. Here, we propose a two-stage spine MRI protocol to localize CSF leak sites in patients with suspected SIH (**Figure 1**). First, 3D HT2-MRM is performed to screen for the presence of SLECs throughout the spine. In the presence of a SLEC or other indeterminate findings, a small field-of-view high resolution CISS sequence, referred to as meso-CISS to reflect its mesoscale resolution, is employed to further characterize these abnormalities and enable precise localization of the CSF leak site. The term mesoscale denotes spatial resolutions on the order of hundreds of micrometers (e.g., 0.1–0.5 mm^3^ isotropic voxel size; acquired meso-CISS resolution: 0.5 mm^3^). We present a series of cases where this approach successfully identified dural tears, demonstrating its potential in enhancing diagnostic accuracy for SIH.

**FIG 1.**
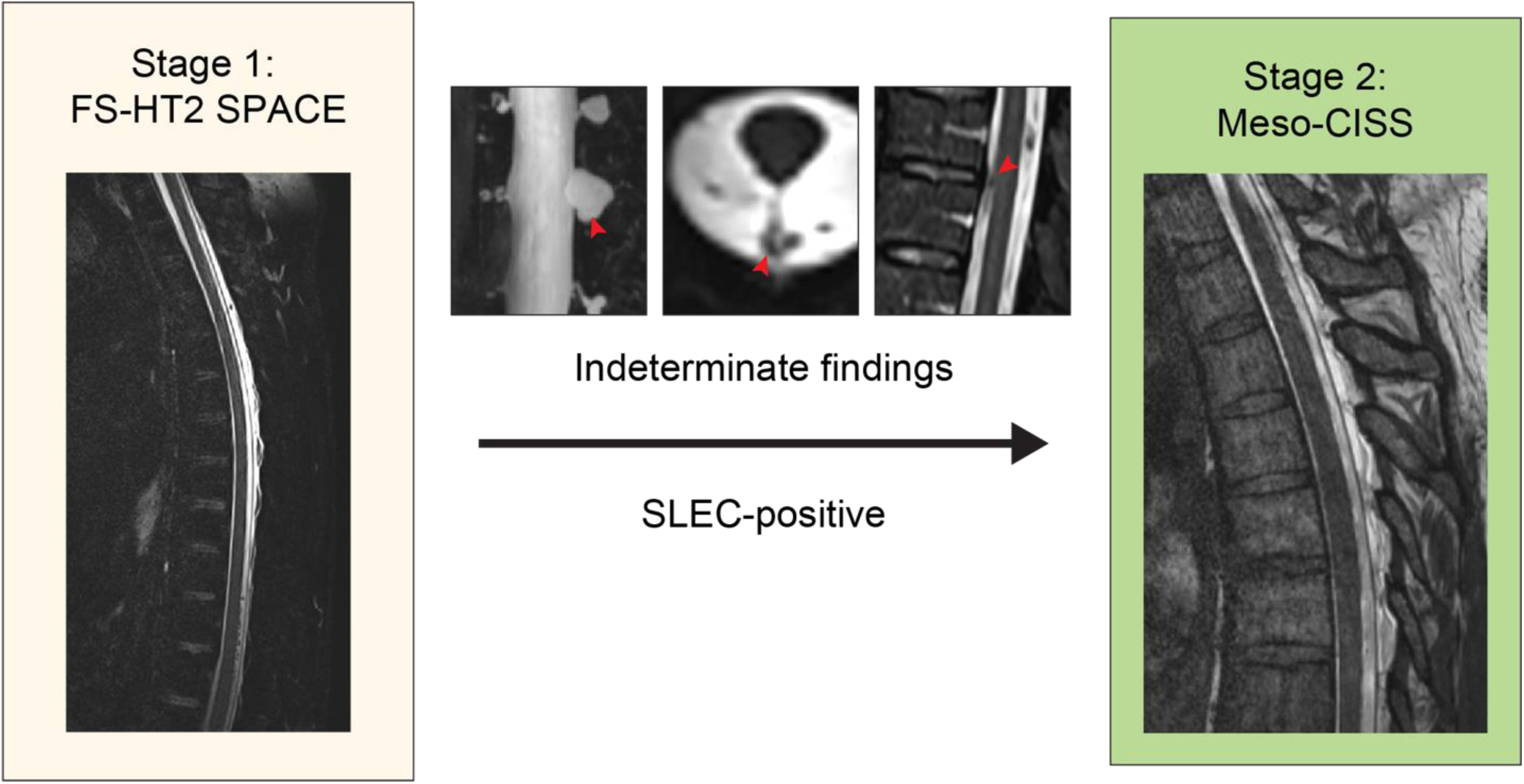
Spine MRI Protocol for CSF Leak Detection. Patients with suspected SIH underwent total spine MRI using an HT2-MRM sequence for SLEC detection. Positive SLEC findings and indeterminate findings such as abnormal perineural cysts, arachnoid scarring, or flow artifacts, were further evaluated with targeted meso-CISS imaging and multiplanar reconstruction to localize dural defects.

## METHODS

### Patient Population

This retrospective study included a total of 20 patients with suspected SIH or CSF leak between May 2024 and July 2025 who underwent total spine MRI with the CSF leak protocol (**Table 1**). Brain MR images were available in 70% of patients and were evaluated for tonsillar ectopia. Bern score was calculated for 60% of patients who had a contrast enhanced brain MRI[24].

**Table 1.** Main Characteristics of the Study Population. Data are reported as number of patients. Mean data are ± SD, with ranges in parenthesis. ^1^ SLEC: spinal longitudinal epidural fluid collection ^2^ SIH: Spontaneous intracranial hypotension

| Characteristic | Patients<br>(*n* = 20) | SLEC<sup>1</sup>-positive patients<br>(*n* = 8) |
| --- | --- | --- |
| Demographic Characteristics |  |  |
| Age (y) – mean ± SD (range) | 49 ± 15 (17-75) | 41 ± 13 (17-58) |
| Sex, Female – N (%) | 15 (75) | 7 (88) |
| Clinical Features – N (%) |  |  |
| Headache | 16 (80) | 8 (100) |
| Neck pain | 7 (35) | 4 (50) |
| Nausea | 6 (30) | 3 (38) |
| Auditory disturbance | 6 (30) | 2 (25) |
| Visual disturbance | 5 (25) | 0 (0) |
| Gait disturbance | 5 (25) | 0 (0) |
| Brain Imaging Findings |  |  |
| Median Bern SIH<sup>2</sup> score (0-9 points) – mean (IQR) | 6 (5) | 8 (2) |
| Cerebellar ectopia – N (%) | 10 (50) | 6 (75) |

### MRI Workflow

This protocol included HT2-MRM sequence optimized for uniform shimming and accelerated high-resolution imaging across cervical, thoracic, and lumbar stations to achieve total spine coverage. HT2-MRM images were reviewed in real time by a neuroradiologist. For patients with a positive SLEC on HT2-MRM, targeted imaging was performed using a limited field-of-view meso-CISS sequence acquired at one to two imaging stations to cover the full extent of the SLEC. For patients with indeterminate findings, such as arachnoid scarring, abnormal perineural cysts, or flow-related artifacts, a single-station meso-CISS acquisition was employed to further evaluate the abnormality (**Figure 1**). Inpatients were scanned on the nearest available 3T unit using the best HT2-MRM parameters available on the scanner. All meso-CISS scans were acquired using a standardized protocol regardless of inpatient or outpatient status. Multiplanar reconstruction was employed to locate dural defects.

### Total Spine MRI CSF Leak Protocol

All MRI scans were performed using 3T Siemens scanners, with all outpatient imaging acquired on a Magnetom Vida. For HT2-MRM, fat saturated sagittal 3D T2 SPACE sequences of the cervical, thoracic and lumbar spine were acquired with a repetition time (TR) of 1800 ms, an echo time (TE) of 221 ms, and a constant flip angle of 100°. The field of view (FOV) was set to 280-320 mm, with an isotropic voxel size of 0.7 x 0.7 x 0.7 mm^3^ and a slice thickness of 0.7 mm. Scan times were as follows: cervical spine, 5:22; thoracic spine, 5:18; and lumbar spine, 4:42. An acceleration factor of 8 was applied using compressed sensing with a bandwidth of 657-789 Hz/pixel. Fat suppression was achieved using SPectral Attenuated Inversion Recovery (SPAIR) with absolute shimming. The sagittal 3D meso-CISS sequences were acquired with a TR of 5.2 ms, a TE of 2.27 ms, and a flip angle of 50°. The FOV was 150 mm, with a voxel size of 0.5 x 0.5 x 0.5 mm^3^, a slice thickness of 0.5 mm, and receiver bandwidth was 460 Hz/pixel. The acquisition time per station was 5 minutes and 45 seconds.

## RESULTS

### Patient Overview

Among the 20 consecutive patients who underwent total spine MRI with the CSF leak protocol for suspected SIH (**Table 1**), 8 (40%) were identified as SLEC-positive on HT2-MRM, warranting further evaluation with meso-CISS imaging to precisely localize potential dural defects. Meso-CISS was performed in an additional 8 patients where other abnormalities, such as scarring, perineural cysts, or diverticula on HT2-MRM required further clarification. Meso-CISS identified findings suspected to represent dural defects in five patients (Patients 1-4, 6), with confirmatory evidence from DSM and/or surgery in two cases. In the fifth patient, an apparent intrathecal protrusion of ligamentum flavum calcification was observed on meso-CISS, likely occurring through a dural defect and corresponding to the contrast leak site identified on a prone CT myelography. In the sixth patient, the initial meso-CISS did not demonstrate a definite dural defect; however, a subsequent meso-CISS revealed a type 1 ventral dural tear. Meso-CISS did not reveal a definite dural defect in the seventh patient with a dorsal SLEC from T2-T9, though evaluation was limited by patient motion. In the eighth SLEC-positive patient, extensive spinal instrumentation caused significant metal artifact on meso-CISS, rendering the study nondiagnostic. Of note, all imaging findings reported here were identified during the initial interpretation of MRI, without the availability of DSM or CTM at the time of review. In all cases, the meso-CISS finding was first identified and subsequently used to guide correlation with the HT2-MRM images.

### Dural Defect Detection

#### Patient 1

The first patient was a female in her 30’s presenting with orthostatic headaches and tinnitus. Brain MRI performed six weeks prior to the dedicated CSF leak protocol demonstrated 5 mm cerebellar tonsillar ectopia and a Bern score of 8, indicating a high likelihood of positive spinal imaging[24, 25]. HT2-MRM imaging revealed a ventral SLEC extending from C4-T10 and a dorsal SLEC at T3-T4. Subsequent meso-CISS imaging localized a ventral dural defect measuring 2 × 8 mm at T8-T9 which communicated with the ventral SLEC, consistent with a type 1 CSF leak (**Figure 2A**). A comparison with the HT2-MRM SPACE sequence (**Figure 2A**) highlights the superior resolution of meso-CISS in localizing the defect to this level. This corresponded to a ‘flow void sign’ on the 2D sagittal STIR imaging (**Figure 2B**). The dural defect location was subsequently confirmed via digital subtraction myelography (**Figure 2C**).

**FIG 2.**
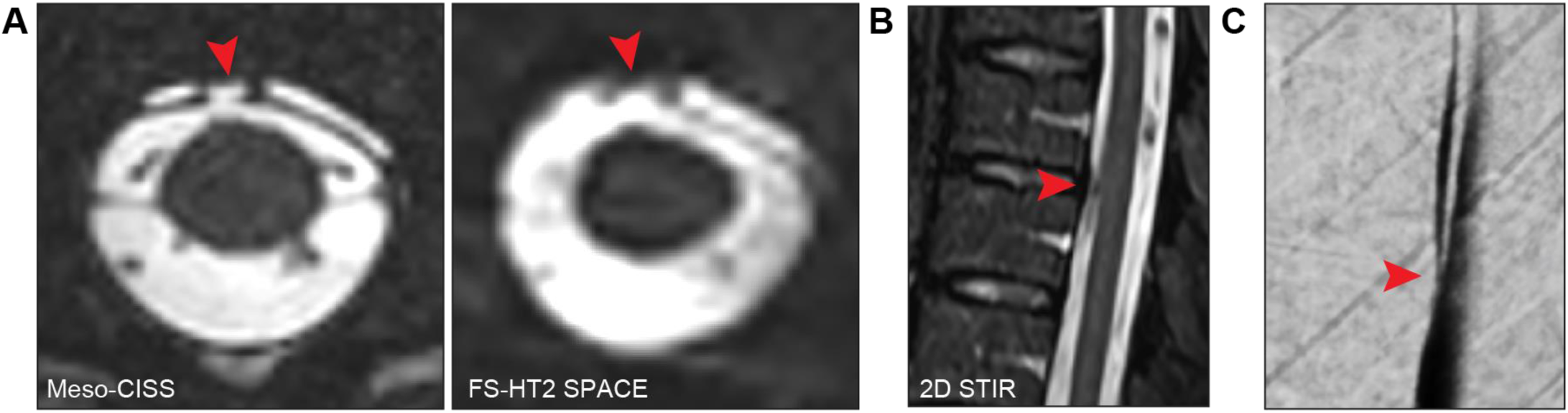
Type 1 CSF Leak with a Ventral Dural Tear. (**A**) Axial meso-CISS (left) and HT2-MRM (right) sequences demonstrating a ventral dural tear near the T8-T9 disc space (red arrowhead). (**B**) Sagittal STIR imaging illustrates the ’flow void sign’ (red arrowhead). (**C**) Digital subtraction myelography confirms a ventral dural defect near the T8-T9 disc space (red arrowhead).

The patient initially underwent an epidural blood patch procedure with injection at T8-L3 followed by Tisseel fibrin sealant application into the dorsal epidural space at T8-T9, but her symptoms persisted. Ultimately, she underwent a left T8-T9 transpedicular partial corpectomy, diskectomy, and duraplasty of a large ventral defect measuring 1-2 cm at the inferior border of T8 with improvement in her symptoms. One month postoperatively, she reported experiencing mild persistent headaches, tinnitus and brain fog. However, approximately three months postoperatively, the patient reported progressively worsening orthostatic headaches. Repeat CTM revealed a persistent ventral CSF leak at the T8-T9 disc space, with a thin ventral epidural fluid collection extending cranially to the level of C2. Notably, the size of the dural defect appeared reduced compared to preoperative imaging. Based on these findings, the patient underwent a left T8-T9 foraminotomy and CSF leak repair. Intraoperatively, a 5 mm ventral dural defect was identified at this level and was repaired using DuraGen, TachoSil, and Tisseel, which were layered over the defect. At her one-month postoperative follow-up, the patient reported improvement in her positional headaches but noted the onset of intermittent, frontal, pressure-like headaches triggered by activity, suggestive of rebound intracranial hypertension. She will continue to follow with the neurosurgery clinic for ongoing assessment and management of her symptoms.

#### Patient 2

The second patient was a female in her 30’s presenting with orthostatic headaches, neck pain, and auditory disturbances. A dedicated brain MRI was not performed. However, total spine imaging demonstrated 8 mm cerebellar tonsillar ectopia. HT2-MRM imaging revealed a long segment SLEC extending from C6-L2, which was circumferential but thickest dorsally, measuring up to 4 mm at T7-T9. The collection tapered below L2 with an additional SLEC extending from L4-L5 to S2. Subsequent meso-CISS imaging localized a focal outpouching at the left T11-T12 neural foramen, corresponding to a meningeal diverticulum with an adjacent dural defect measuring 3 x 3 mm (**Figure 3**). DSM and post-myelography spot images demonstrated a type 2 CSF leak from the left T11-T12 meningeal diverticulum (**Figure 3**). The patient did not undergo treatment. Two months after the DSM, she reported spontaneous resolution of symptoms.

**FIG 3.**
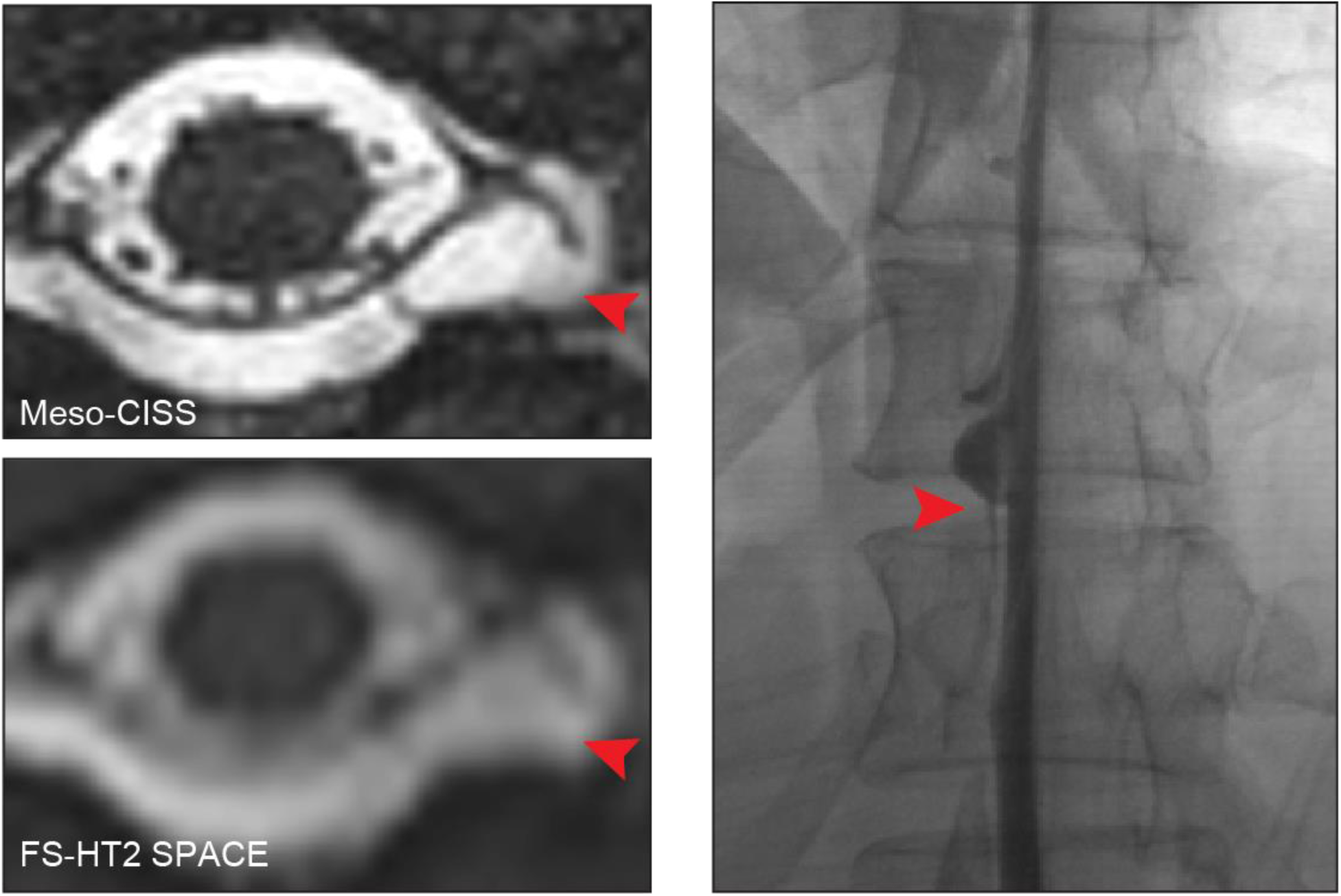
Type 2 CSF Leak from a Left-Sided Meningeal Diverticulum. Axial reconstruction of meso-CISS (top left) and corresponding HT2-MRM imaging (bottom left) localize a focal outpouching at the left T11-T12 neural foramen, consistent with a meningeal diverticulum. The associated dural defect is more clearly visualized on meso-CISS (red arrowhead). Digital subtraction myelography (right) demonstrates linear epidural contrast adjacent to the left T11-T12 neural foramen (red arrowhead).

#### Patient 3

The third patient was a female in her 40’s who presented with non-positional morning headaches and bilateral upper extremity tingling. Brain MRI obtained one week prior to the dedicated CSF leak protocol demonstrated 5 mm of cerebellar ectopia and a Bern score of 6. HT2-MRM imaging revealed both dorsal and ventral SLECs extending from the cervical spine to the upper lumbar spine. Meso-CISS imaging identified a dural defect measuring 2.5 × 1 mm at the tip of a right-sided perineural cyst at T8-T9 (**Figure 4A**), consistent with a type 2 CSF leak. HT2-MRM imaging (**Figure 4B**) demonstrated similar findings retrospectively, though with lower spatial resolution. These corresponded to a “jester shoe” configuration (pointy appearance with a rounded outpouching at the tip) on HT2-MRM maximum intensity projection (MIP) (**Figure 4C**). Unfortunately, the patient was lost to follow-up.

**FIG 4.**
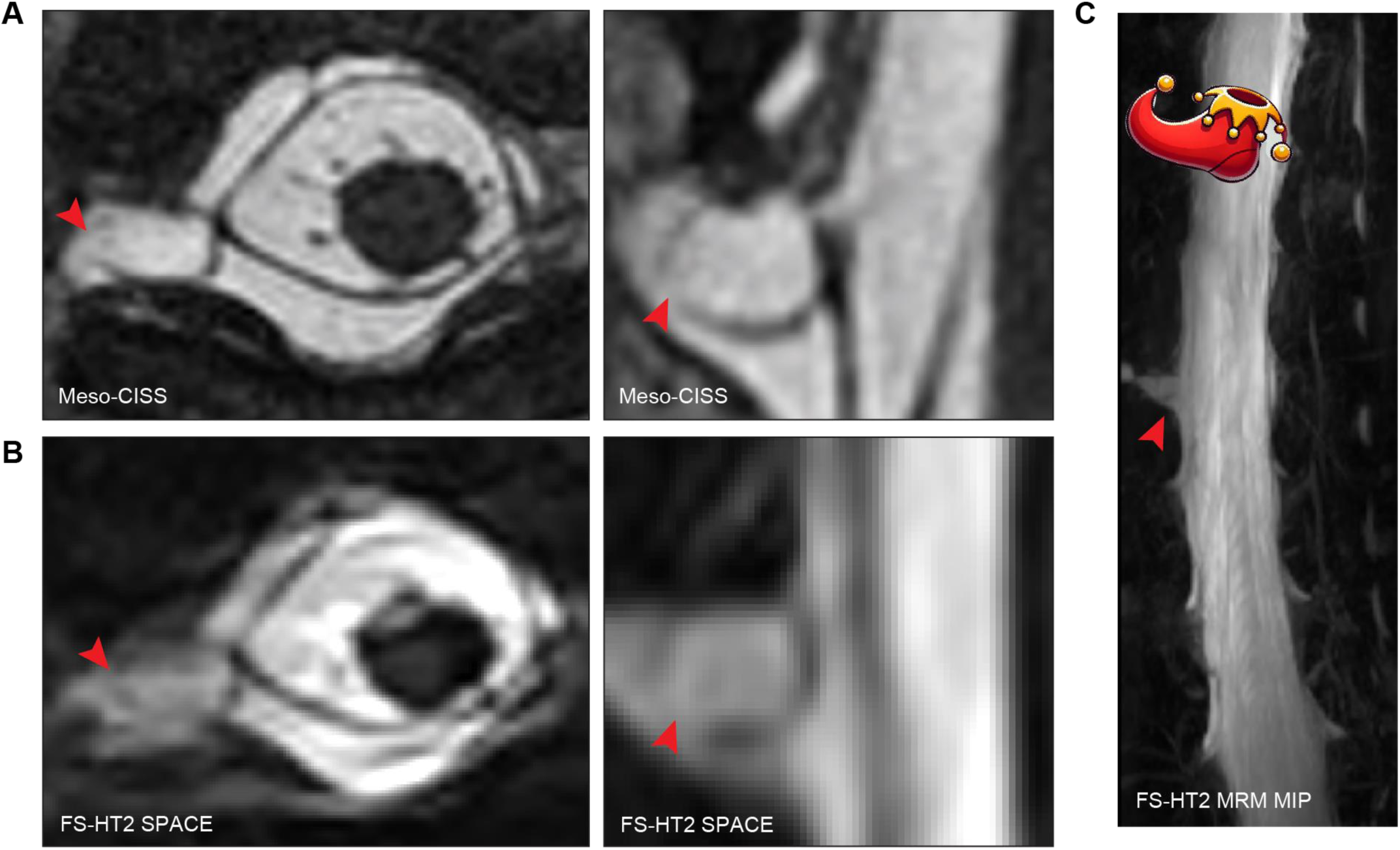
Type 2 CSF Leak from a Right-Sided Perineural Cyst. (**A**) Meso-CISS and (**B**) FS-HT2 SPACE sequences demonstrate a dural defect at the tip of a right-sided perineural cyst at T8-T9 (red arrowhead). (**C**) HT2-MRM maximum intensity projection (MIP) imaging illustrates the corresponding jester shoe configuration.

#### Patient 4

The fourth patient was a female in her 50’s presenting with orthostatic headaches, neck pain, and nausea. Brain MRI performed one week prior to the dedicated CSF leak protocol demonstrated a Bern score of 6. HT2-MRM imaging revealed a large dorsal SLEC extending from C2 along the length of the spinal canal. The 1.5 × 1 mm dural defect was localized to the shoulder of an irregular right nerve root sleeve at T11-T12 on high-resolution imaging (**Figure 5**). This defect communicated with the dorsal SLEC, consistent with a shoulder variant of type 2 CSF leak. The patient underwent two epidural blood patch procedures with temporary symptomatic improvement. She is receiving follow-up care for her persistent headaches at a facility within her insurance network.

**FIG 5.**
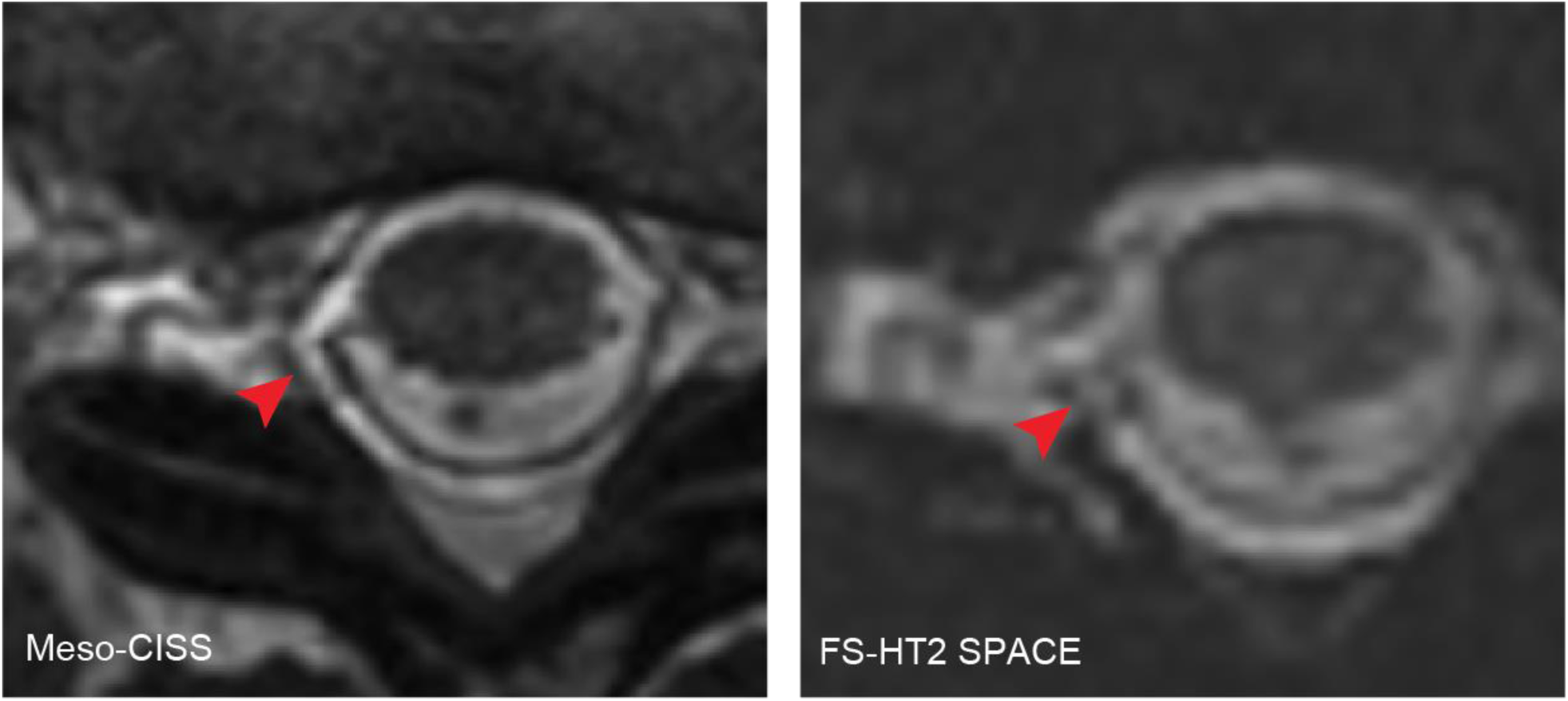
Type 2 CSF Leak from an Irregular Right Nerve Root Sleeve. Axial meso-CISS (left) and HT2-MRM (right) sequences demonstrate a dural defect at the shoulder of an irregular right nerve root sleeve at T11-T12 (red arrowhead), in communication with a large dorsal SLEC.

#### Patient 5

The fifth patient was a female in her 50’s with a history of a prior CSF leak in 2022, successfully treated with an epidural blood patch. She presented with recurrent orthostatic headaches and neck pain. Brain MRI revealed a Bern score of 8. HT2-MRM imaging identified a ventral and dorsal SLEC extending from T3 to L4. Although a dural defect was not directly visualized, meso-CISS imaging demonstrated focal dural wrinkling adjacent to an intrathecal protrusion of the right ligamentum flavum spur at the level of the right T3 facet (**Figure 6**). The patient underwent an epidural blood patch at the T2-T3 interspace the following day. However, one month after the procedure, she reported continued headaches and neck pain, similar in quality and severity to her pre-procedure symptoms. Subsequent CTM demonstrated ossification of the ligamentum flavum and facet arthropathy at the right T3-4 level, with an adjacent posterolateral extra-axial CSF collection. The supine images were similar to the meso-CISS findings, showing the valve-like protrusion of the osseous spur. On prone imaging, the CSF leak through a dorsal dural defect was evident, suggesting a valve-like mechanism of CSF egress related to the ligamentum flavum ossification (**Figure 6**). At her follow-up appointment one month later, the patient reported substantial symptomatic improvement, and surgical intervention was deferred.

**FIG 6.**
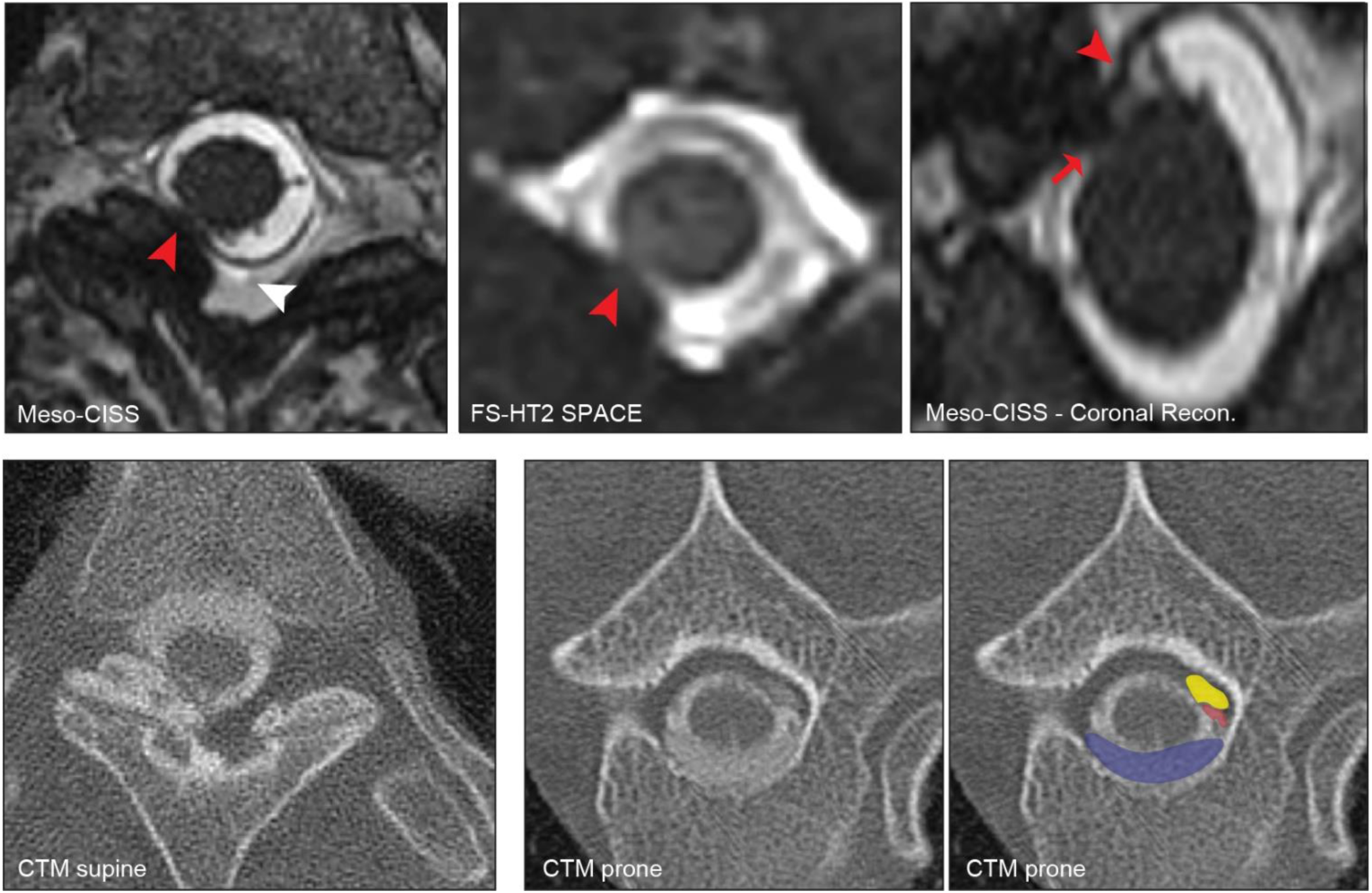
Type 1 CSF Leak from a Ligamentum Flavum Spur. Axial meso-CISS (top left), axial HT2-MRM (top middle), and coronal meso-CISS (top right) imaging showing intrathecal protrusion of a ligamentum flavum spur (red arrow) at the level of the right T3 facet resulting in dural wrinkling (red arrowhead) and CSF leak (white arrowhead). Supine CT myelogram (bottom left) demonstrates the valve-like protrusion of the osseous spur, while prone CT myelogram (bottom right) demonstrates the CSF leak through the dorsal dural defect (red). The osseous spur is highlighted in yellow, and the ventral SLEC is highlighted in blue.

#### Patient 6

The sixth patient was a female in her 40’s who presented with non-positional headaches. Brain MRI findings included a Bern score of 9 and cerebellar ectopia measuring 4 mm. HT2-MRM revealed a predominantly dorsolateral SLEC with trace ventral involvement throughout the thoracic spine. The dorsal collection became asymmetric in the upper thoracic spine towards the right, with a second thickened membranous structure, consistent with neodura (**Figure 7A**)[14]. High resolution imaging identified multiple small defects within the neodura spanning T2-T8. CTM demonstrated predominantly dorsal epidural contrast extending from T3-4 to T8-9 with an initially non-enhancing space (likely a neospace; **Figure 7B**). DSM revealed a non-localizing ventral leak (**Figure 7C**). However, meso-CISS was valuable in identifying neodural membranes, which are strongly associated with ventral leaks[14]. Their presence suggested an underlying ventral dural defect, supporting the diagnosis of a type 1 CSF leak even in the absence of direct visualization. The patient underwent two nontargeted lumbar epidural blood patches at the L2-3 interspace one month apart without improvement in her symptoms. The patient subsequently underwent a repeat CTM which demonstrated a large osteophyte at T5-6 with an adjacent epidural fluid collection extending into the right neural foramen, potentially representing the site of CSF leak. One month later, a repeat total spine MRI using the CSF leak protocol demonstrated a small ventral dural defect at T5-6, partially obscured by a calcified disc spur, consistent with a type 1 ventral leak (**Figure 7D**). Given these findings, the patient is scheduled to undergo a right T5-6 transforaminal discectomy and surgical CSF leak repair.

**FIG 7.**
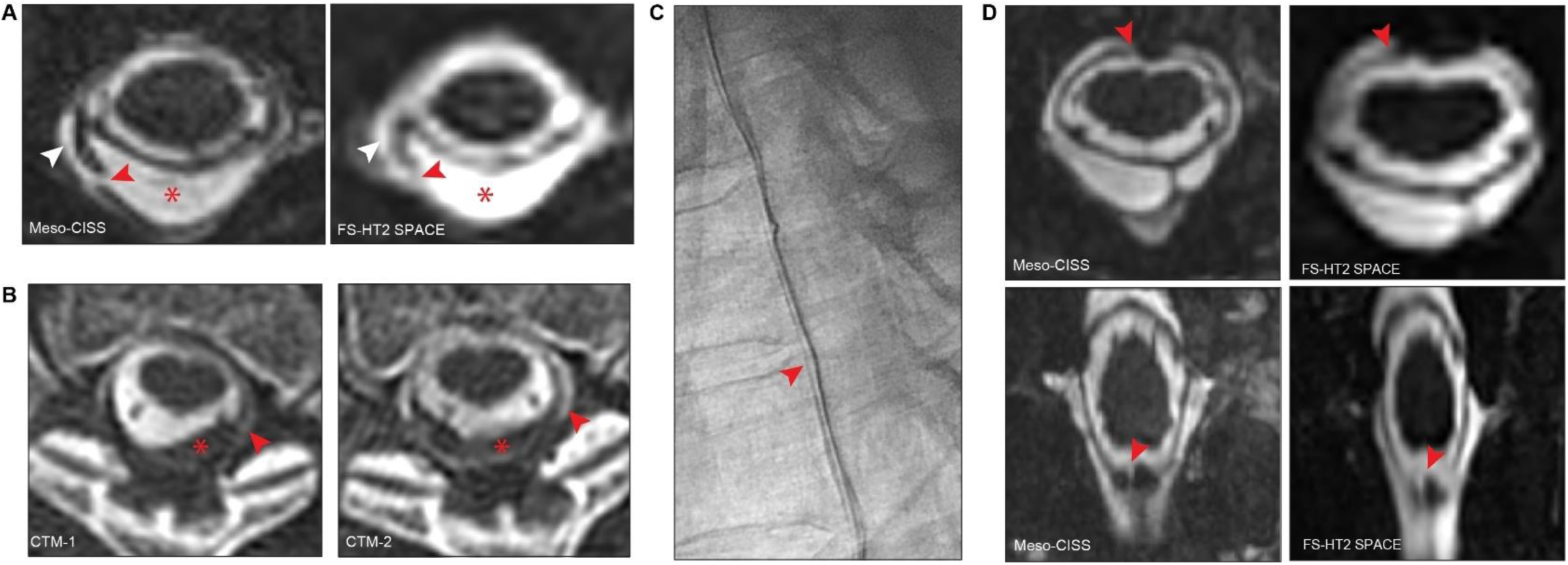
Type 1 CSF Leak with Dorsal Neodura. (**A**) Axial meso-CISS (left) and HT2-MRM (right) imaging showing dural splitting with a defect in the external layer of neodura (red arrowhead) in communication with a right dorsolateral SLEC (white arrowhead) and neospace (asterisk) at the level of T4. (**B**) CT myelograms (CTMs) of the thoracic spine demonstrating a dorsal epidural contrast collection (red arrowhead) and neospace (asterisk). Delayed CTM (CTM-2) shows progressive subtle contrast enhancement in the neospace. (**C**) DSM revealing a non-localizing ventral leak. (**D**) Axial (top) and coronal (bottom) meso-CISS (left) and HT2-MRM (right) images from a repeat spine MRI demonstrating a discontinuity of the ventral dura (red arrowhead) and dural thickening along the right lateral aspect of a calcified T5-T6 disc.

### Ancillary Findings

In addition to its role in localizing CSF leaks, meso-CISS also enhances the visualization of ancillary spinal abnormalities that may provide further diagnostic insight. Arachnoid scarring/adhesions, with or without cord distortion or tethering, were present in 3 patients with available meso-CISS imaging (15%, **Supplemental Figure 1**). Prominent perineural cysts were identified in a total of 13 patients (**Supplemental Figure 2**) and were better delineated on meso-CISS compared to HT2-MRM due to higher spatial resolution. Additionally, meso-CISS exhibits greater robustness to flow artifacts, aiding in distinguishing true findings on HT2-MRM from potential artifacts.

### Limitations

Despite its high spatial resolution, meso-CISS imaging has several limitations, including susceptibility to patient motion and metallic instrumentation (**Supplemental Figure 3**). Another limitation is the small FOV of meso-CISS, which, while enabling high spatial resolution, restricts spinal coverage.

## DISCUSSION

Here, we show that a two-stage spine MRI protocol, combining HT2-MRM and meso-CISS, can be utilized to localize dural defects in SIH. Although diagnostic yields have improved with advanced myelographic techniques, definitive leak localization remains challenging in a subset of cases[26, 27]. Dynamic CTM, in particular, is associated with substantial radiation exposure and may confer an increased risk of radiation-induced malignancy, especially in younger patients or those requiring repeated imaging[28, 29]. These risks, combined with the invasive nature of both dynamic DSM and CTM, have limited the application of these approaches in patients with chronic symptoms or atypical clinical presentations[30]. Therefore, MRI techniques such as meso-CISS, which localize CSF leaks or narrow the field of view for subsequent targeted DSM or CTM may reduce radiation exposure, improve diagnostic efficiency, and, in some cases, eliminate the need for invasive procedures.

Prior studies have reported the use of b-SSFP sequences for detecting spinal dural defects in the context of CSF leak[19–21]. Whole-spine b-SSFP, used alongside CTM, has been shown to aid in localizing ventral dural defects and guiding targeted epidural blood patching[21]. In another study of patients with classical infratentorial superficial siderosis, b-SSFP sequences were used to localize ventral dural defects and guide surgical repair[20]. Expanding on these earlier efforts, this small-scale study of a two-stage spine MRI workflow integrates HT2-MRM for detecting SLECs and targeted meso-CISS for precise CSF leak localization. Notably, in all five patients with findings suggestive of dural defects on meso-CISS, the suspected defects were prospectively identified during the initial MRI interpretation prior to any confirmatory myelography. In each case, the corresponding abnormality on HT2-MRM was either subtle or not recognized until retrospective review, prompted by the meso-CISS findings.

Furthermore, this imaging strategy proved effective in localizing not only ventral type 1 leaks, but also a broader spectrum of type 2 leaks. In all cases, dural defects were smaller than 3 mm in at least one dimension, highlighting the value of the sharpness and isotropic resolution of meso-CISS images.

Recent studies suggest that indirect MRI findings may aid in localizing CSF leaks[11, 12]. For instance, a recent study reported that a ‘flow void sign’ on 2D turbo spin-echo T2-weighted imaging co-localizes with the site of a ventral dural tear[11]. While these findings could be subtle and are not definitive, we observed a similar flow void co-localizing with a ventral dural defect in Patient 1, suggesting that flow void sign may serve as an adjunct to HT2-MRM for positioning meso-CISS and CSF leak localization. Similarly, the ‘bud-on-branch’ sign has been associated with type 2 lateral leaks. However, these signs may appear at multiple levels beyond the site of the CSF leak. Additionally, in Patient 3, we noted a ‘jester shoe’ appearance of a perineural cyst with dural defect at the tip, representing a previously uncharacterized variant of a type 2 CSF leak. Taken together, these observations highlight that while indirect MRI signs are nonspecific, they may serve as valuable adjunctive markers to SLECs for optimizing meso-CISS field-of-view positioning and refining CSF leak localization.

Beyond subtle indirect MRI signs, meso-CISS may also identify structural abnormalities that contribute to dural defect localization. Although a dural defect was not directly visualized in Patient 5, meso-CISS delineated the intrathecal protrusion of a calcified ligamentum flavum, likely through a dural defect. This finding was associated with a dorsal SLEC and wrinkling of the adjacent dura. These findings suggest that meso-CISS may serve as a valuable tool for identifying and characterizing intrathecal osseous protrusions (e.g., endplate spurs, calcified intervertebral discs, and calcified ligamentum flavum), which may partially seal and obscure an underlying dural defect.

As exemplified by Patient 6, meso-CISS also demonstrated value in depicting complex pathology, including formation of neodura and neospace. These vascularized neomembranes, which typically arise dorsally and often coexist with ventral leaks, may mimic native dura intraoperatively, potentially complicating surgical dissection and increasing the risk of hemorrhage[14, 31]. In this case, meso-CISS provided high-resolution visualization of the external neo-membrane, differentiating neodural remodeling from primary dura and revealing multiple discrete neodural defects with greater spatial resolution than HT2-MRM. As such, meso-CISS may aid preoperative planning by identifying these membranes and guiding surgical approach. Similarly, meso-CISS demonstrated superior sensitivity to arachnoid scarring compared with HT2-MRM.

In addition to its diagnostic utility, the meso-CISS technique may offer value in longitudinal assessment of CSF leaks. While resolution of epidural fluid remains the primary marker of leak closure, meso-CISS may offer complementary structural assessment of defects over time, particularly in patients undergoing conservative management or post-intervention follow-up. Furthermore, this technique may support improved investigation of the natural history of spinal dural CSF leaks, offering insights into their temporal evolution and the durability of various treatment approaches.

This study is limited by its small sample size. Further studies across multiple centers are warranted to further validate the clinical utility of the two-stage spine MRI protocol incorporating HT2-MRM and meso-CISS for CSF leak localization. Additionally, while meso-CISS provides high-resolution isotropic imaging, its performance is susceptible to motion artifacts. As a result, patient motion can degrade image quality, potentially limiting its diagnostic value in patients who have difficulty remaining still. Furthermore, meso-CISS is largely non-diagnostic in the presence of extensive metallic instrumentation, as susceptibility artifacts can obscure fine structural details and preclude accurate leak detection. Although high spatial resolution was achieved with meso-CISS in this study (0.5 mm^3^), the dural defects identified in this study were often smaller than 2 mm in at least one dimension, underscoring the need for even higher-resolution imaging to enhance the precision of CSF leak detection. Ultra-high-field MRI offers a promising approach to improving signal-to-noise ratio and imaging resolution. However, higher field strengths exacerbate susceptibility-induced signal inhomogeneities and B1+ field nonuniformity, which degrade and limit spine imaging quality[32]. Furthermore, banding artifacts in b-SSFP imaging become increasingly pronounced at ultra-high fields[33, 34]. Advances in coil design, optimized shimming techniques[35], and alternative sequence strategies are essential to overcoming these challenges while leveraging the improved signal-to-noise ratio for more precise localization of spinal CSF leaks.

The results of this study support the use of meso-CISS as a targeted, high-resolution imaging tool to refine CSF leak localization. In our cohort, meso-CISS enabled prospective identification of suspected dural defects that were only retrospectively appreciated on HT2-MRM. By improving pre-procedural localization, this approach has the potential to increase the diagnostic yield and spatial accuracy of invasive studies, such as DSM and CTM, reduce unnecessary radiation exposure, and facilitate more precise and targeted therapeutic interventions. Further studies across multiple centers are warranted to refine these imaging approaches and evaluate the broader clinical impact of incorporating meso-CISS into the diagnostic algorithm for SIH, including its potential role in monitoring the progression or resolution of CSF leaks under different management strategies.

## Data Availability

All data produced in the present study are available upon reasonable request to the authors.

## ACKNOWLEDGEMENT

We thank Melissa Affolter, Chris Harrison, Amanda Woelfel, and Jzan Karla Modesto for their critical role in implementing the CSF leak imaging protocol and ensuring protocol adherence and appropriate patient workup throughout the clinical workflow.

