## Supplemental Figures 1-3 for "Mesoscale CISS Imaging for the Detection of Dural Defects in Spinal CSF Leaks"

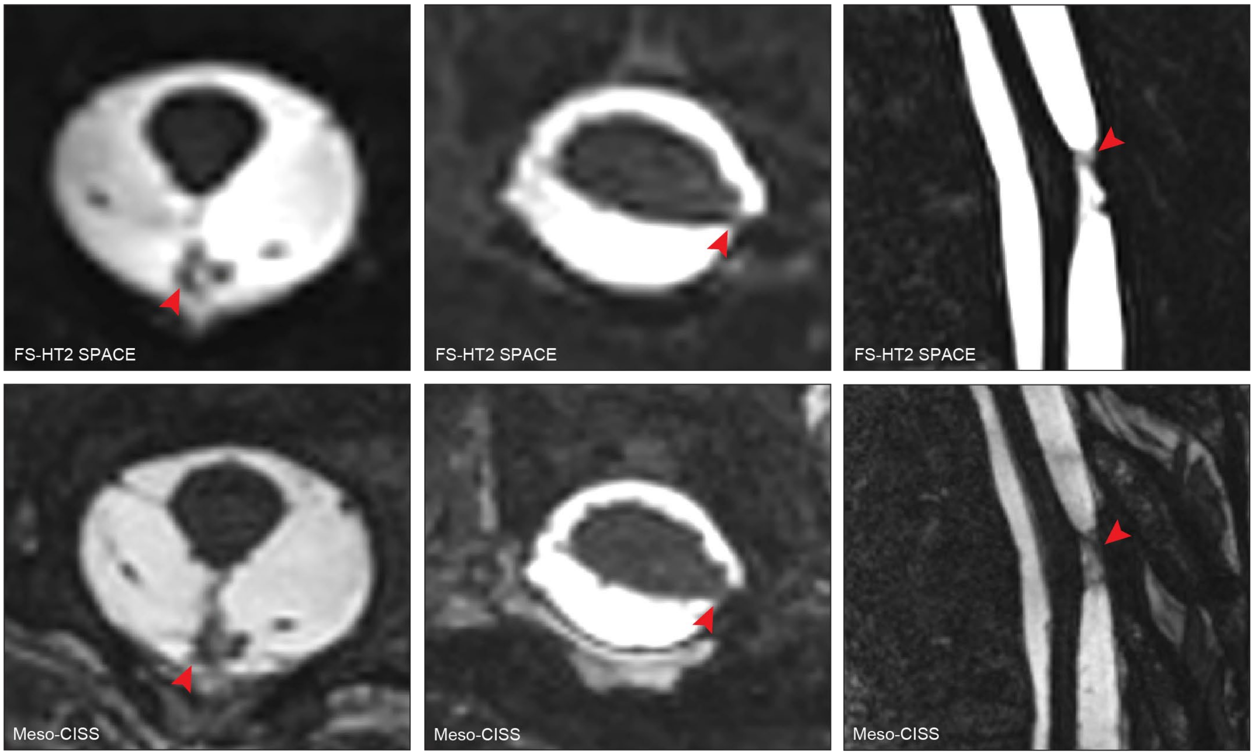


**Supplemental Figure 1.** Arachnoid Scarring. (Left) Axial meso-CISS and HT2-MRM imaging demonstrates scarring and thickening of the median dorsal septum with mild deformity of the dorsal cord at the level of T7. (Middle) Axial meso-CISS and HT2-MRM imaging showing mild eccentric leftward cord tethering and arachnoid scarring (red arrowhead) at a site of prior left T6-T7 laminectomy. (Right) Sagittal meso-CISS and HT2-MRM imaging demonstrate thin adhesions with dorsal tethering of the spinal cord at T6-T7 (red arrowhead).

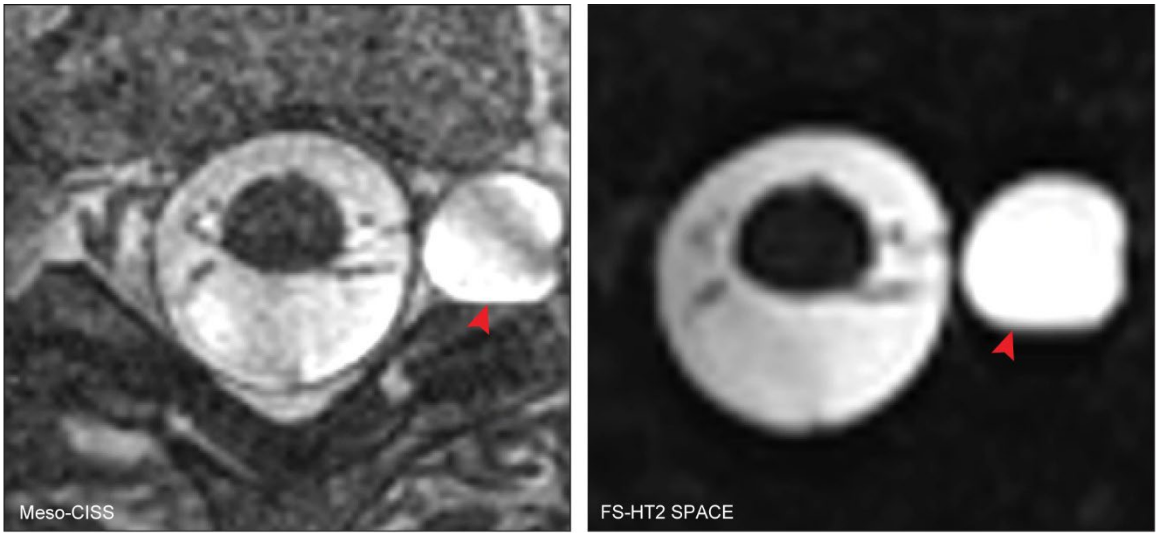


**Supplemental Figure 2.** Axial meso-CISS (left) and FS-HT2 SPACE imaging (right) showing a left-sided perineural cyst at the level of T4.

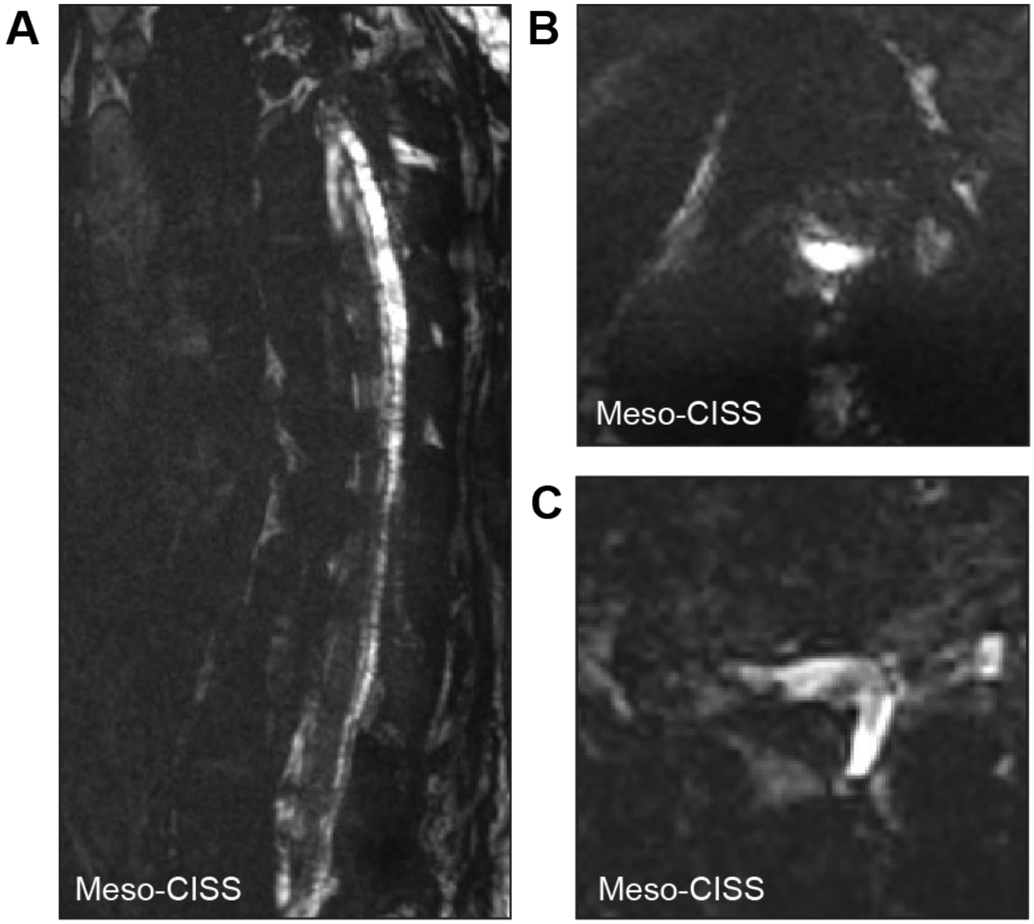


**Supplemental Figure 3.** (**A**) Sagittal and (**B, C**) axial meso-CISS images depicting obscuration of the thoracic spinal cord in patients with posterior instrumented spinal fusion.
